# Assessing the burden of COVID-19 in Canada

**DOI:** 10.1101/2020.06.14.20130815

**Authors:** Simona Bignami-Van Assche, Ari Van Assche

**Author notes:** Funding: None. Competing interests: None.

## Abstract

**Background:** The burden of COVID-19 in Canada is unequally distributed geographically, with the largest number of cases and fatalities recorded in Québec and Ontario while other provinces experienced limited outbreaks. To date, however, no study has assessed how provincial epidemics have unfolded in a comparative perspective. This is essential to calibrate projections of the future course of the epidemic and plan health care resources for the second wave of infections.

**Methods:** Using newly released individual-level data collected by the Public Health Agency of Canada, we assess COVID-19-related morbidity and mortality across age and gender groups at the provincial level through a combination of demographic and survival analyses.

**Results:** Québec has the highest absolute and per capita number of COVID-19 confirmed positive cases, hospitalizations and fatalities in all age groups. In each province, a higher number of women than men test positive for the disease, especially above age 80. Yet consistently across age groups, infected men are more likely to be hospitalized and enter intensive care than women do. These gender differences in hospitalisation rates account for the higher case fatality risk due to COVID-19 among men compared to women.

**Interpretation:** Although health care capacity across provinces has been sufficient to treat severe cases, we find that the main factor accounting for gender differences in COVID-19-related mortality is the need for hospitalization and intensive care, especially above age 80. This suggests a selection effect of severe cases requiring to be treated in a hospital setting that needs to be further investigated.

## INTRODUCTION

The new coronavirus disease, first identified in Wuhan, China, in December 2019, has now spread around the globe infecting almost seven million people, and causing more than 400,000 deaths [*1*]. In Canada, the first two cases of COVID-19 were confirmed in a couple returning to Ontario from Wuhan on January 23, 2020. Travellers returning to Québec and Alberta from abroad after spring break ignited transmission in these two provinces as well. As of May 31, 90,516 positive cases and 7,092 deaths of COVID-19 were confirmed nationwide [*2*].

The burden of COVID-19 is unequally distributed across provinces in Canada [*2*]. Québec and Ontario account for more than 80 percent of these cases (approximately 50 and 30 percent respectively) and 95 percent of deaths. At the other end of the spectrum, the Atlantic provinces have been the least affected by COVID-19 and recorded less than 2 percent of the total number of confirmed positive cases and less than one percent of deaths. The Northwest Territories, Yukon, and Nunavut closed their borders after a handful of cases had been identified and did not experience community transmission of COVID-19.

Research about the burden of COVID-19 in Canada at the provincial level has, so far, been limited. The spread (cumulative number of cases and deaths) and intensity (daily counts of reported cases and deaths) of COVID-19 across provinces are accessible via provincial health agencies and several online dashboards, notably the one maintained by the COVID-19 Canada Open Data Working Group described in a recent publication of *CMAJ* [*3*]. Studies have focused on severe outcomes of the disease in specific settings [*4*], overall fatality [*5*], and excess mortality in Québec [*6*]. Epidemiological models of the future course of the epidemic have also been developed at the provincial and federal levels to plan for health care resources’ allocation [*7-11*]. Yet, there is no comparative evidence about how provincial epidemics actually unfolded in terms of disease severity and health care utilization across age and gender groups. In this paper, we fill this gap and we assess the burden of COVID-19 across provinces in Canada.

## METHODS

### Data sources

The analysis exploits an individual-level file compiled by Statistics Canada from information provided by the Public Health Agency of Canada since late March [*12*].^1^ On May 24, this file was updated to include several variables on COVID-19 confirmed positive patients’ trajectory, from infection to hospitalization and death or recovery.

As of its latest update, which refers to the period until May 31, the dataset includes information on 90-100% of cases and fatalities recorded at the provincial level, with the exception of British Columbia (see Annex Table 1). Information on the age and gender characteristics of confirmed positive cases is 99% complete in all provinces. For fatalities, age or gender information is missing for less than 1 percent of cases in Québec, Ontario and the Atlantic provinces; 4 percent of cases in the Prairie provinces; and 16 percent of cases in British Columbia. The dataset also includes individual-level data for 70 to 100% of confirmed positives cases who were hospitalized or admitted to intensive care in each province, with information on their age and gender missing in less than 4 percent of cases across provinces. Overall, for approximately a third of COVID-19 confirmed positive cases in each province information is missing about individuals’ clinical status after testing (hospitalisation, death and recovery). This phenomenon, which has been highlighted in Europe as well [*13*], is related to the natural evolution of the infection over time as well as to delays in reporting. Nonetheless, it is not evenly distributed across age groups: in Québec, clinical status is missing for approximately a third of confirmed positive individuals age 80+, compared to 5 percent in Ontario. In addition, for 60 percent of cases with missing clinical status, COVID-19 diagnosis had occurred in the previous 4 or more weeks, that is, well after the average survival time for the disease had elapsed [*5*].

**Table 1.** Ratios of COVID-19 confirmed positive cases, deaths and hospitalizations per 100,000 people, by age group and province, as of May 31

| Age group | National<sup>1</sup> | Québec<sup>1</sup> | Ontario<sup>1</sup> | Prairies<sup>1</sup> | B.C.<sup>2</sup> | Atlantic<sup>1</sup> |
| --- | --- | --- | --- | --- | --- | --- |
| **Cases** |  |  |  |  |  |  |
| Total | 228 | 534 | 200 | 104 | 51 | 60 |
| 0-19 | 49 | 104 | 36 | 50 | 8 | 27 |
| 20-39 | 219 | 512 | 192 | 123 | 51 | 61 |
| 40-59 | 265 | 604 | 232 | 149 | 65 | 61 |
| 60-79 | 210 | 440 | 200 | 74 | 51 | 63 |
| 80+ | 1024 | 2592 | 814 | 141 | 143 | 165 |
| **Deaths** |  |  |  |  |  |  |
| Total | 19 | 52 | 16 | 2 | 3 | 2 |
| 0-19 | 0 | 0 | 0 | 0 | 0 | 0 |
| 20-39 | 0 | 0 | 0 | 0 | 0 | 0 |
| 40-59 | 2 | 3 | 2 | 0 | 0 | 0 |
| 60-79 | 24 | 58 | 22 | 4 | 4 | 3 |
| 80+ | 313 | 809 | 251 | 44 | 48 | 29 |
| **Hospitalizations** |  |  |  |  |  |  |
| Total | 23 | 50 | 25 | 5 | 7 | 3 |
| 0-19 | 1 | 3 | 1 | 1 | 0 | 0 |
| 20-39 | 6 | 13 | 6 | 2 | 2 | 0 |
| 40-59 | 18 | 33 | 21 | 6 | 5 | 3 |
| 60-79 | 44 | 83 | 49 | 12 | 14 | 5 |
| 80+ | 180 | 409 | 168 | 26 | 33 | 12 |

Sources: <sup>1</sup> [12]. <sup>2</sup> [14].
Notes: The individual-level file compiled by Statistics Canada from information supplied by PHAC groups provinces in three regions: Ontario; Prairies and Northwest Territories; British Columbia and Yukon; Atlantic provinces New Brunswick, Nova Scotia, Prince Edward Island, Newfoundland and Labrador). Hospitalizations include cases that required intensive care.

**Table 2.** Percentage of COVID-19 confirmed positive cases who were hospitalized, and percentage of hospitalized cases who received intensive care as of May 31, by gender and province

|  | Both sexes |  | Men |  | Women |  |
| --- | --- | --- | --- | --- | --- | --- |
|  | Hospitalized | ICU | Hospitalized | ICU | Hospitalized | ICU |
| National | 10.2 | 19.4 | 12.3 | 23.2 | 8.7 | 15.5 |
| Québec | 9.4 | 17.6 | 11.4 | 21.2 | 8.2 | 14.3 |
| Ontario | 12.4 | 21.7 | 15.1 | 25.9 | 10.4 | 16.9 |
| Prairies | 5.1 | 8.7 | 5.7 | 9.8 | 4.4 | 6.6 |
| B.C. | 17.3 | 29.5 | 22.9 | 31.9 | 12.5 | 26.1 |
| Atlantic provinces | 5.1 | 16.9 | 6.2 | 16.2 | 2.9 | 16.0 |

Source: Our calculations from [12].

### Statistical analysis

First, we describe the magnitude of provincial COVID-19 epidemics in Canada by comparing confirmed positive cases, mortality, and hospitalization ratios per capita across provinces. For British Columbia, due to the low coverage of COVID-19-related deaths in Statistics Canada’s dataset, we draw from information disseminated directly by the provincial Center for Disease Control [*14*].

Second, we describe COVID-19 severity across age and gender groups in each province with: a) demographic pyramids of confirmed positive, hospitalized, recovered, and fatal cases; and b) estimates of age- and sex-specific rates of severe outcomes (hospitalization among all confirmed positive cases, and intensive care admission among hospitalized cases).

Finally, we estimate the case fatality risk (CFR), or the proportion of confirmed cases who result in fatalities, by hospitalization status. The severity profile of a novel pathogen is one of the most critical issues as it begins to spread, when assessing disease course and outcome is crucial for planning health interventions [*5, 15*]. For this reason, the CFR is an important epidemiological indicator to monitor during the current outbreak of COVID-19. With our individual-level dataset, the CFR can be correctly estimated through event history modelling to take into account censoring that arises because, at the time of observation, the outcome is unknown for a nonnegligible portion of infected individuals. In this framework, the CFR coincides with the cumulative incidence (CI) of COVID-19-related mortality, with recovery as a competing risk.^2^ The CI and its standard error are estimated controlling for gender and hospitalization status with *stcrreg* in Stata/SE (version 12.0, StataCorp, LLC).

### Ethics approval

This research was based on publicly available data and did not require ethics approval.

## RESULTS

### Case, mortality and hospitalization ratios

Nationally, 228 positive cases per 100,000 were confirmed in Canada (Table 1). Québec and Ontario have recorded the highest absolute number of COVID-19 cases, and also the highest number relative to population size (534 and 200 per 100,000, respectively). The Prairie provinces have had half as many infections per capita (104 per 100,000, mainly concentrated in Alberta) than Ontario has. British Columbia and the Atlantic provinces have a similar ratio of cases per capita (60 and 51 per 100,000, respectively). As for confirmed positive infections, mortality ratios are highest in Québec at all ages, and particularly at age 80+. In the age group 60-79, the number of deaths per capita in Québec are more than twice as high as in Ontario (58 vs 22 per 100,000), but approximately the same as in the other provinces.

Above age 80, death rates in Québec are more than three times those recorded in Ontario (809 vs 251 per 100,000), and approximately 4 per 100,000 in the other provinces. Finally, Québec has the highest ratio of hospitalizations for all age groups. Under age 40, hospitalization ratios in Québec are more than twice than in Ontario, and between age 40 and 79 years they are 1.5 times higher. Particularly striking is the comparison at age 80+, where the hospitalization ratio is 409 per 100,000 in Québec, 168 per 100,000 in Ontario and 12 per 100,000 in the Atlantic provinces.

### Age-sex pyramids

Figure 1 shows that, with the exception of the Prairies, the age and gender distribution of confirmed positive cases is remarkably similar across provinces. A higher number of women than men are confirmed positive with COVID-19 at all ages, but the number of men who are hospitalized for the disease and admitted to intensive care is higher than for women. The main exception is the age group above 80 years, where a larger number of women test positive and are hospitalized for COVID-19 than men. The burden of mortality for COVID-19 is carried by the elderly. Fatalities are concentrated above age 50 for both men and women and the largest number of COVID-19 confirmed positive cases who died is found among those age 80+ years across all provinces, especially for women.^3^ Nonetheless, a higher number of women than men recovers from the disease at all ages, including above 80 years.

**Figure 1.**
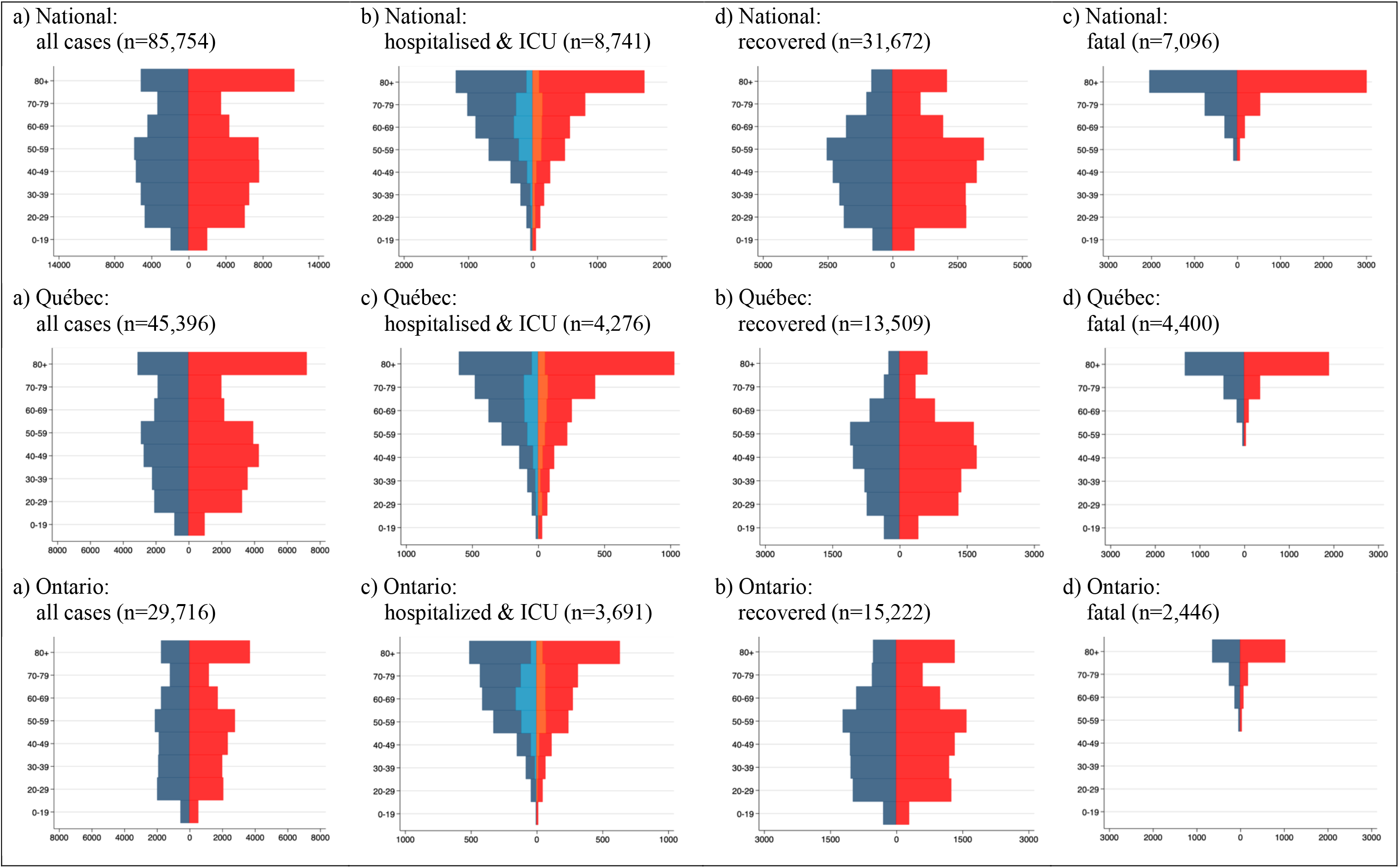
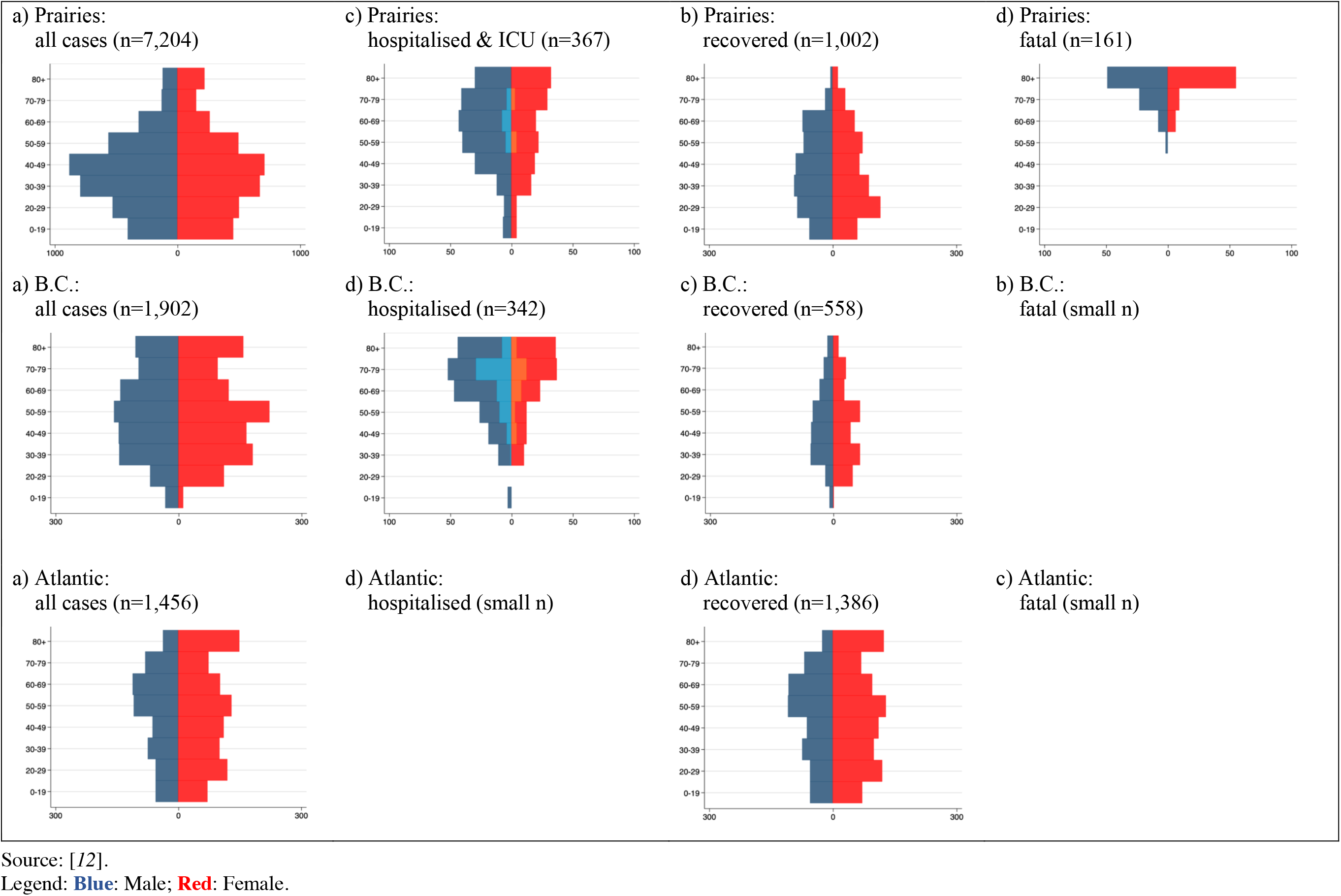
Age-sex pyramids of COVID-19 confirmed positive, hospitalised, fatal and recovered cases as of May 31, by province

### Age- and sex-specific rates of severe outcomes

Nationally, 10.2 percent of COVID-19 confirmed positive cases were hospitalized for the disease. Of these, 19.4 percent needed intensive care. The lowest proportion of hospitalized cases is found in the Atlantic and Prairie provinces (4.5 and 5.1 percent, respectively). British Columbia has recorded the highest proportion of hospitalized cases (17.3 percent) as well as of those who needed intensive care among them (29.5 percent). Compared to men, in all provinces a lower proportion of COVID-19-positive women needed hospitalization and intensive care because of the disease.

The probability of hospitalisation increases sharply with age for both men and women (Figure 2). The reduced hospitalization rate among the oldest age group is related to the fact the most infections at age 80+ in Canada (notably in Québec, Ontario and British Columbia) have occurred in elderly homes. The decreasing probability of severe hospitalization with age is a pattern observed in many countries, and it is suggested to result from clinical decisions about the use of ventilator capacity in most severe cases [*13*].

**Figure 2.**
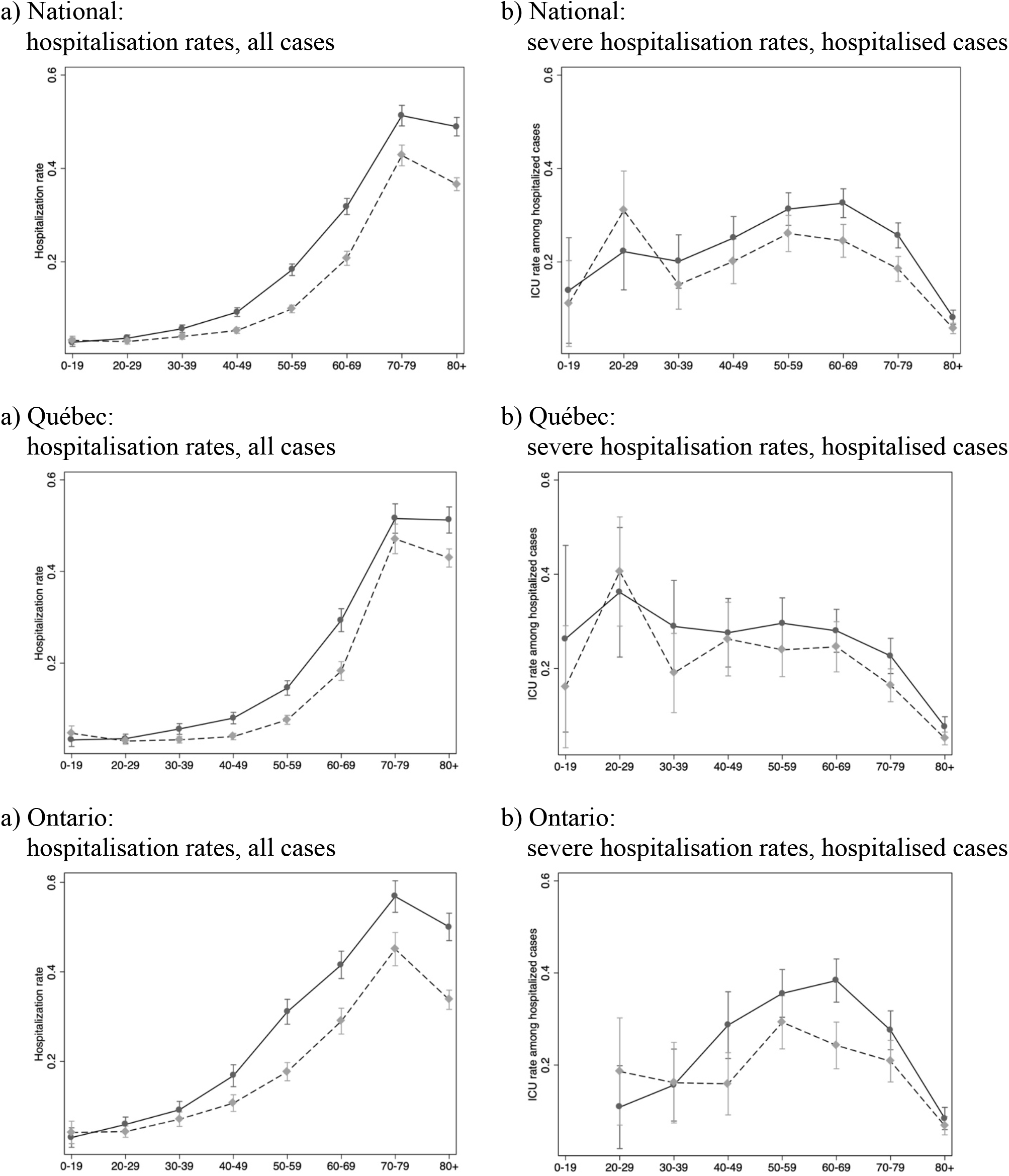
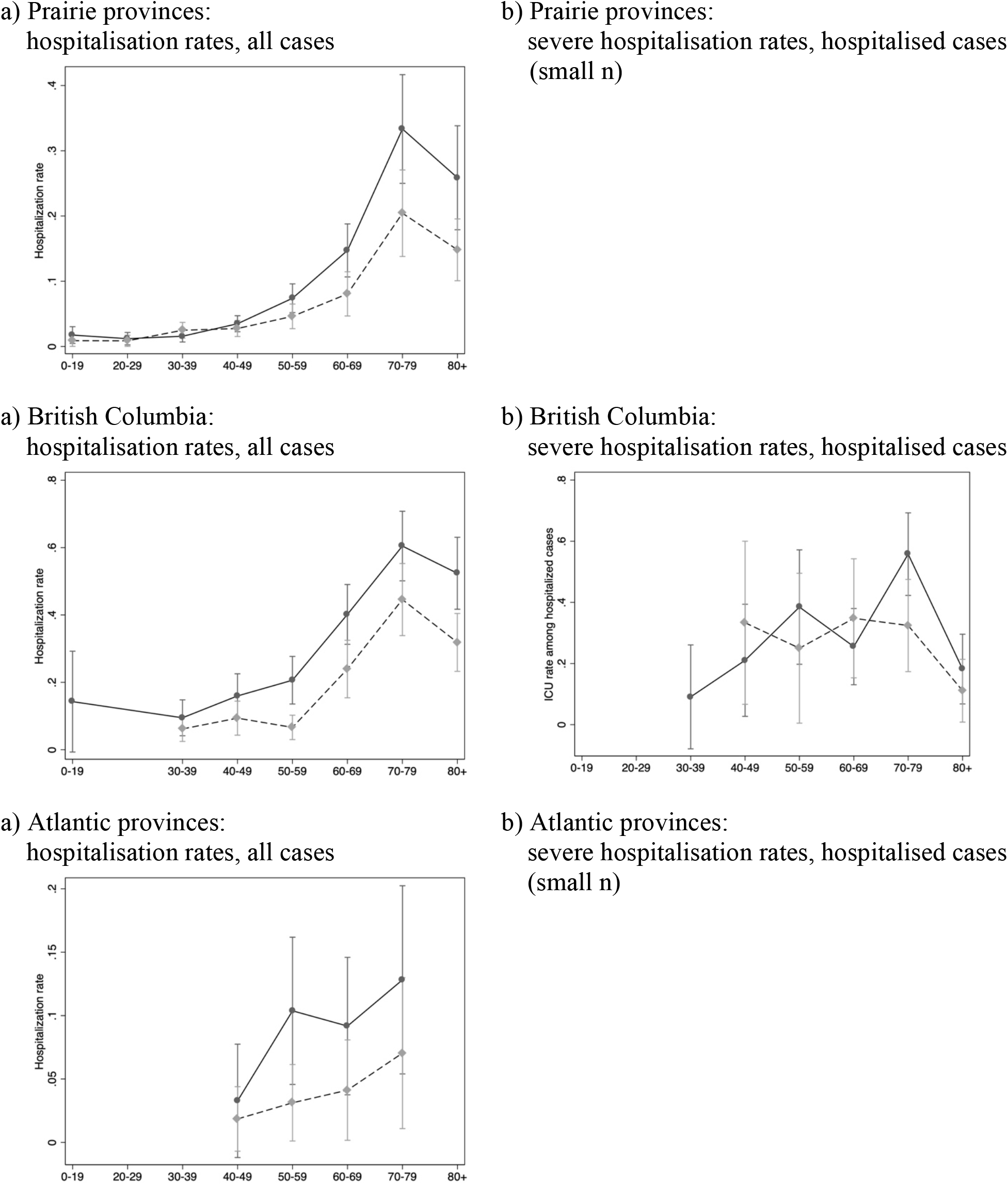
Age- and gender-specific rates of hospitalization among all confirmed positive cases, and rates of admission to intensive care among hospitalized patients (95% confidence intervals in parentheses), by province Source: Our calculations from [*12*].

### Case fatality risk

Table 3 presents our estimates of the CFR across age groups and provinces by adjusting for gender and hospitalization, with or without admission to intensive care. Men have a consistently higher risk of dying for COVID-19 than women do. Nonetheless, men and women who did not need to be hospitalized or admitted to intensive care because of COVID-19 have a very similar risk of dying at all ages, the main exception being men age 80+. Hospitalization indeed increases the mortality risk by 3-4 times, and admission to intensive care by more than 6 times. National-level estimates match closely those for Québec, where the majority of COVID-19 related deaths and hospitalizations have occurred.

**Table 3.** CFR (cumulative incidence of mortality vs recovery) for COVID-19 confirmed positive cases as of May 31, controlling for gender and hospitalization status, by province

| Age group | Men |  |  |  | Women |  |  |  |
| --- | --- | --- | --- | --- | --- | --- | --- | --- |
|  | Total | ICU | Hosp. | Non-hosp. | Total | ICU | Hosp. | Non-hosp. |
| NATIONAL |  |  |  |  |  |  |  |  |
| 50-59 | 0.2 | 1.1 | 0.6 | 0.2 | 0.2 | 1.0 | 0.5 | 0.1 |
| 60-69 | 1.0 | 5.0 | 2.9 | 0.7 | 0.9 | 4.3 | 2.5 | 0.7 |
| 70-79 | 3.2 | 15.6 | 9.2 | 2.5 | 2.7 | 13.4 | 7.8 | 2.1 |
| 80+ | 11.9 | 48.3 | 31.2 | 9.4 | 10.1 | 42.8 | 27.2 | 8.0 |
| All ages | 4.1 | 17.6 | 11.1 | 3.3 | 3.5 | 15.4 | 9.6 | 2.8 |
| QUÉBEC |  |  |  |  |  |  |  |  |
| 50-59 | 0.2 | 0.7 | 0.5 | 0.1 | 0.1 | 0.5 | 0.4 | 0.1 |
| 60-69 | 0.9 | 3.7 | 2.6 | 0.7 | 0.7 | 2.7 | 2.0 | 0.5 |
| 70-79 | 3.5 | 13.7 | 10.0 | 2.8 | 2.6 | 10.3 | 7.5 | 2.1 |
| 80+ | 13.8 | 45.7 | 35.5 | 11.3 | 10.3 | 36.1 | 27.5 | 8.4 |
| All ages | 4.6 | 15.7 | 12.0 | 3.7 | 3.4 | 12.3 | 9.3 | 2.8 |
| ONTARIO |  |  |  |  |  |  |  |  |
| 50-59 | 0.3 | 1.7 | 0.8 | 0.2 | 0.3 | 1.7 | 0.8 | 0.2 |
| 60-69 | 1.2 | 6.8 | 3.3 | 0.8 | 1.2 | 6.7 | 3.2 | 0.8 |
| 70-79 | 3.1 | 17.2 | 8.4 | 2.1 | 3.1 | 16.9 | 8.3 | 2.1 |
| 80+ | 10.4 | 49.4 | 26.0 | 7.1 | 10.2 | 46.9 | 25.7 | 7.0 |
| All ages | 4.3 | 20.6 | 11.0 | 3.0 | 4.3 | 20.4 | 10.1 | 2.9 |
| REST OF CANADA |  |  |  |  |  |  |  |  |
| 50-59 | 0.1 | 0.3 | 0.2 | 0.1 | 0.1 | 0.2 | 0.2 | 0.1 |
| 60-69 | 0.6 | 2.2 | 1.8 | 0.5 | 0.5 | 1.8 | 1.5 | 0.4 |
| 70-79 | 2.5 | 8.7 | 7.3 | 2.2 | 2.0 | 7.1 | 5.9 | 1.8 |
| 80+ | 11.0 | 34.8 | 29.9 | 10.1 | 8.9 | 29.1 | 24.8 | 8.2 |
| All ages | 2.6 | 8.5 | 7.3 | 2.4 | 2.1 | 7.1 | 6.0 | 1.9 |

Source: our calculations from [12].

## INTERPRETATION

As the first wave of the COVID-19 pandemic in Canada is coming to an end, we provide the first comparative assessment of how it unfolded at the provincial level across age and gender groups.

Nationally, the number of COVID-19 infections per capita (252 per 100,000) is similar to that recorded in France [*19*]. Yet, ranking above Belgium and Sweden, Québec has the highest absolute and per capita number of COVID-19 confirmed positive cases as well as hospitalizations and fatalities in all age groups. Age differentials in infection across provinces can be partly explained by different testing strategies for COVID-19. Until the end of March, Québec and Ontario primarily tested symptomatic individuals for COVID-19, whereas in April their governments focused testing on health care workers and elderly patients in the attempt to contains outbreaks in nursing homes that had already infected a large proportion of residents and staff. Alberta, on the contrary, tested health care workers and nursing home personnel in early March when there were few confirmed positive cases in health facilities, and then expanded testing to all symptomatic individuals and, recently, asymptomatic ones as well.

Consistent with evidence from European countries [*13, 20*], a higher number of women than men test positive for the disease in all provinces, but men are more likely to be hospitalized, enter intensive care, and die for the disease than women do. In Canada, women 80+ years old are an exception because they are more likely than men to have severe outcomes.

The need for intensive care has been one of the most important public health issues in dealing with the COVID-19 pandemic. Indeed, efforts aimed at ‘flattening the curve’ of infections, in Canada and around the world, meant to ensure that health care capacity was sufficient to treat severe cases in need of intensive care [*21*]. We find that, nationally, 19.2 percent of hospitalized cases were admitted to intensive care, which is almost twice as high as the European average (11 percent), although the proportion of cases that needed to be hospitalized in Canada is much lower (10.2 percent compared to 35 percent).

Our estimates about men’s higher risk of dying for COVID-19 at all ages compared to women are in line with earlier studies in hospital settings [*20, 22-24*].^4^ An important contribution of our study is to identify that, at the population level, there is no gender difference in the CFR for individuals that do not need to be hospitalized. Once hospitalization is needed, however, the CFR is higher for men across age groups, especially above age 80. This suggests a selection effect of severe cases requiring to be treated in an hospital setting that needs to be further investigated. Specifically, information on the duration of hospitalization before death or recovery, as well as on patients’ presentation and pre-existing conditions, is essential to explore the mechanisms behind our findings [*20, 22-24*]. Unfortunately, the Public Health Agency of Canada has no immediate plan to release these data (personal communication).

In spite of this limitation, our results about disease severity and utilization of hospital resources for COVID-19 are important to calibrate epidemiological projections and appropriately plan health care resources for the second wave of infections expected in the fall.

## Data Availability

Data is freely available.

https://www150.statcan.gc.ca/t1/tbl1/en/tv.action?pid=1310078101

## ACKNOWLEDGEMENTS

We would like to acknowledge that Daniela Ghio, at the Joint Research Center European Commission, participated to all stages of manuscript development and data analysis. We thank Sylvan Tremblay at Statistics Canada and Claire Crawford at the Public Health Agency of Canada for helpful discussions about the data. We also would like to thank participants to the seminar series of the Center for Population Dynamics at McGill University for their comments and questions on an earlier version of this manuscript.

---

**ANNEX Table 1.** Official count of the cumulative number of COVID-19 confirmed positive cases, deaths, recoveries and hospitalizations and coverage of the individual-level file compiled by Statistics Canada from information shared by PHAC as of May 24, by province

|  | National | Québec | Ontario | Prairies | Atlantic | B.C. |
| --- | --- | --- | --- | --- | --- | --- |
| **Cases** |  |  |  |  |  |  |
| Official count<sup>1</sup> | 90,516 | 50,651 | 27,859 | 7,936 | 1,473 | 2,584 |
| In StatCan's file<sup>2</sup> | 85,754 | 45,396 | 29,716 | 7,204 | 1,456 | 1,982 |
|  | (83%) | (91%) | (100%) | (91%) | (99%) | (78%) |
| *Age/gender missing* | 483 (.56%) | 140 (.31%) | 287 (.97%) | 17<br>(.24%) | 15<br>(1.0%) | 24<br>(1.2%) |
| **Deaths** |  |  |  |  |  |  |
| Official count<sup>1</sup> | 7,092 | 4,439 | 2,266 | 161 | 63 | 164 |
| In StatCan's file<sup>2</sup> | 7,096 | 4,400 | 2,446 | 161 | 52 | 37 |
|  | (99%) | (98%) | (100%) | (100%) | (83%) | (24%) |
| *Age/gender missing* | 112 (1.6%) | 29<br>(.66%) | 65<br>(2.7%) | 9<br>(5.6%) | 2<br>(3.9%) | 7<br>(18.9%) |
| **Hospitalizations** |  |  |  |  |  |  |
| Official count<sup>3</sup> | n/a | 3,933 | 3,262 | n/a | n/a | 488 |
| In StatCan's file<sup>2</sup> | 8,741 | 4,276 | 3,691 | 367 | 65 | 342 |
|  | (n/a) | (98%) | (99%) | (n/a) | (n/a) | (99%) |
| *Age/gender missing* | 41<br>(2.4%) | 6<br>(.80%) | 20<br>(2.5%) | 8<br>(25.0%) | 1<br>(9.1%) | 6<br>(5.9%) |
| **Recoveries** |  |  |  |  |  |  |
| Official count<sup>1</sup> | 48,573 | 16,070 | 21,810 | 7,103 | 1,386 | 2,191 |
| In StatCan's file<sup>2</sup> | 31,672 | 13,509 | 15,222 | 1,002 | 1,386 | 558 |
|  | (59%) | (85%) | (54%) | (14%) | (100%) | (27%) |
| *Age/gender missing* | 134 (.42%) | 12<br>(.09%) | 92<br>(.60%) | 8<br>(.80%) | 11<br>(.79%) | 11<br>(2.0%) |

Sources: <sup>1</sup> [4]. <sup>2</sup> [12]. <sup>3</sup> Québec [12]; Ontario [20]; British Columbia [21]; Atlantic and Prairies provinces [12].
Note: The individual-level file compiled by Statistics Canada from information supplied by PHAC groups provinces in three regions: Ontario; Prairies (Alberta, Manitoba and Saskatchewan) and Northwest Territories; British Columbia and Yukon; Atlantic provinces. Hospitalizations include cases that required admission to intensive care.

**ANNEX Figure 1.**
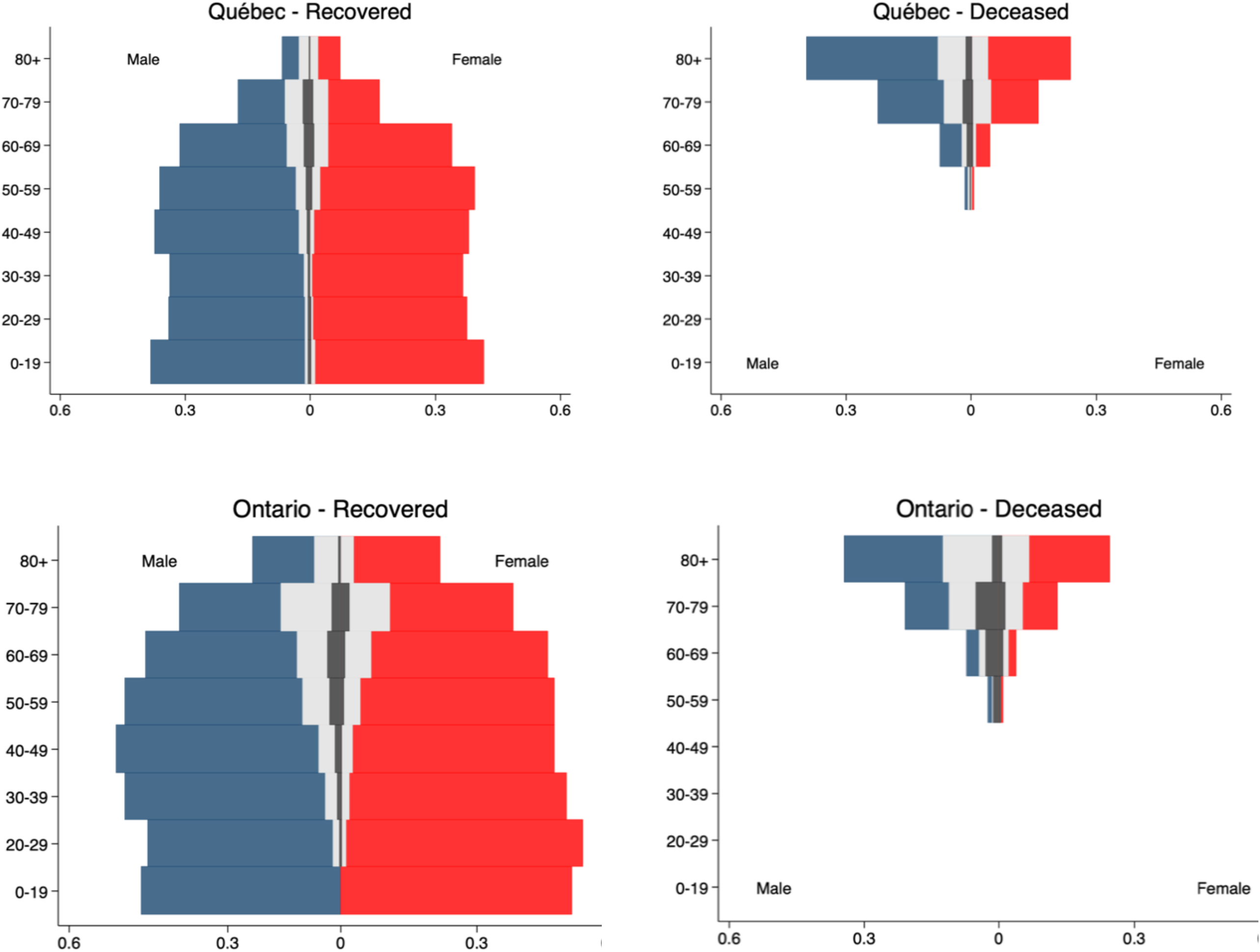
Age and gender distribution of the proportion of COVID-19 confirmed positive cases in Québec and Ontario who recovered (to the left) and who died (to the right) as of May 24, by whether they were hospitalized (in light grey) or in intensive care (in dark grey)

## Footnotes

1 The governments of Alberta and Ontario disseminate an individual-level file containing information on the age and gender of each COVID-19 confirmed positive case. This file covers all individual cases confirmed in each province but does not include information on individuals’ hospitalization status.

2 When calculated from aggregate data on confirmed cases and deaths, the CFR is a simple ratio between the former and the latter, which is prone to numerous biases [*5*; *15*-*17*]. With individual-level data, the CFR can be estimated as a true measure of risk as the proportion of incidence for the disease [*18*].

3 Above age 70, the largest proportion of cases resulting in fatalities had been hospitalized or needed intensive care for the disease (see Annex Figure 1).

4 A recent study in *CMAJ* [*5*] estimated that the CFR for COVID-19 confirmed positive cases in Canada as of April 22 was 5.5% (CI: 4.9 – 6.4%) when adjusting the observed CFR for the delay between disease onset and death. Our estimates at the national level for men and women (Table 2) are lower than this value because we control for hospitalization status and, as indicated earlier, for a sizable proportion of cases the clinical outcome (death or recovery) is still unrecorded. Adjustments of the observed CFR thus need to take into account this source of bias in addition to distortions introduced by confirmed positive cases’ survival time.

